# Integrating endometrial proteomic and single cell transcriptomic pipelines reveals distinct menstrual cycle and endometriosis-associated molecular profiles

**DOI:** 10.1101/2022.01.29.22269829

**Authors:** Lauren Baugh, Brittany A. Goods, Juan S. Gnecco, Yunbeen Bae, Michael Retchin, Constantine N. Tzouanas, Megan Loring, Keith Isaacson, Alex K. Shalek, Douglas Lauffenburger, Linda Griffith

**Affiliations:** Center for Gynepathology Research, Massachusetts Institute of Technology, Cambridge, MA; Biological Engineering Department, Massachusetts Institute of Technology, Cambridge, MA; Thayer School of Engineering at Dartmouth College, Hanover, NH; Department of Chemistry, Institute for Medical Engineering & Science, and Koch Institute for Integrative Cancer Research Massachusetts Institute of Technology, Cambridge, MA; Division of Health Science & Technology, Harvard Medical School, Cambridge, MA; Broad Institute of MIT and Harvard, Cambridge, MA; Ragon Institute of MGH, MIT, and Harvard, Cambridge, MA; Newton Wellesley Hospital, Newton, MA

## Abstract

Endometriosis is a debilitating gynecological disorder affecting approximately 10% of the female population. Despite its prevalence, robust methods to classify and treat endometriosis remain elusive. Changes throughout the menstrual cycle in tissue size, architecture, cellular composition, and individual cell phenotypes make it extraordinarily challenging to identify markers or cell types associated with uterine pathologies since disease-state alterations in gene and protein expression are convoluted with cycle phase variations. Here, we developed an integrated workflow to generate both proteomic and single-cell RNA-sequencing (scRNA-seq) data sets using tissues and cells isolated from the uteri of control and endometriotic donors. Using a linear mixed effect model (LMM), we identified proteins associated with cycle stage and disease, revealing a set of genes that drive separation across these two biological variables. Further, we analyzed our scRNA-seq data to identify cell types expressing cycle and disease-associated genes identified in our proteomic data. A module scoring approach was used to identify cell types driving the enrichment of certain biological pathways, revealing several pathways of interest across different cell subpopulations. Finally, we identified ligand-receptor pairs including Axl/Tyro3 – Gas6, that may modulate interactions between endometrial macrophages and/or endometrial stromal/epithelial cells. Analysis of these signaling pathways in an independent cohort of endometrial biopsies revealed a significant decrease in Tyro3 expression in patients with endometriosis compared to controls, both transcriptionally and through histological staining. This measured decrease in Tryo3 in patients with disease could serve as a novel diagnostic biomarker or treatment avenue for patients with endometriosis. Taken together, this integrated approach provides a framework for integrating LMMs, proteomic and RNA-seq data to deconvolve the complexities of complex uterine diseases and identify novel genes and pathways underlying endometriosis.

**Graphical abstract:** 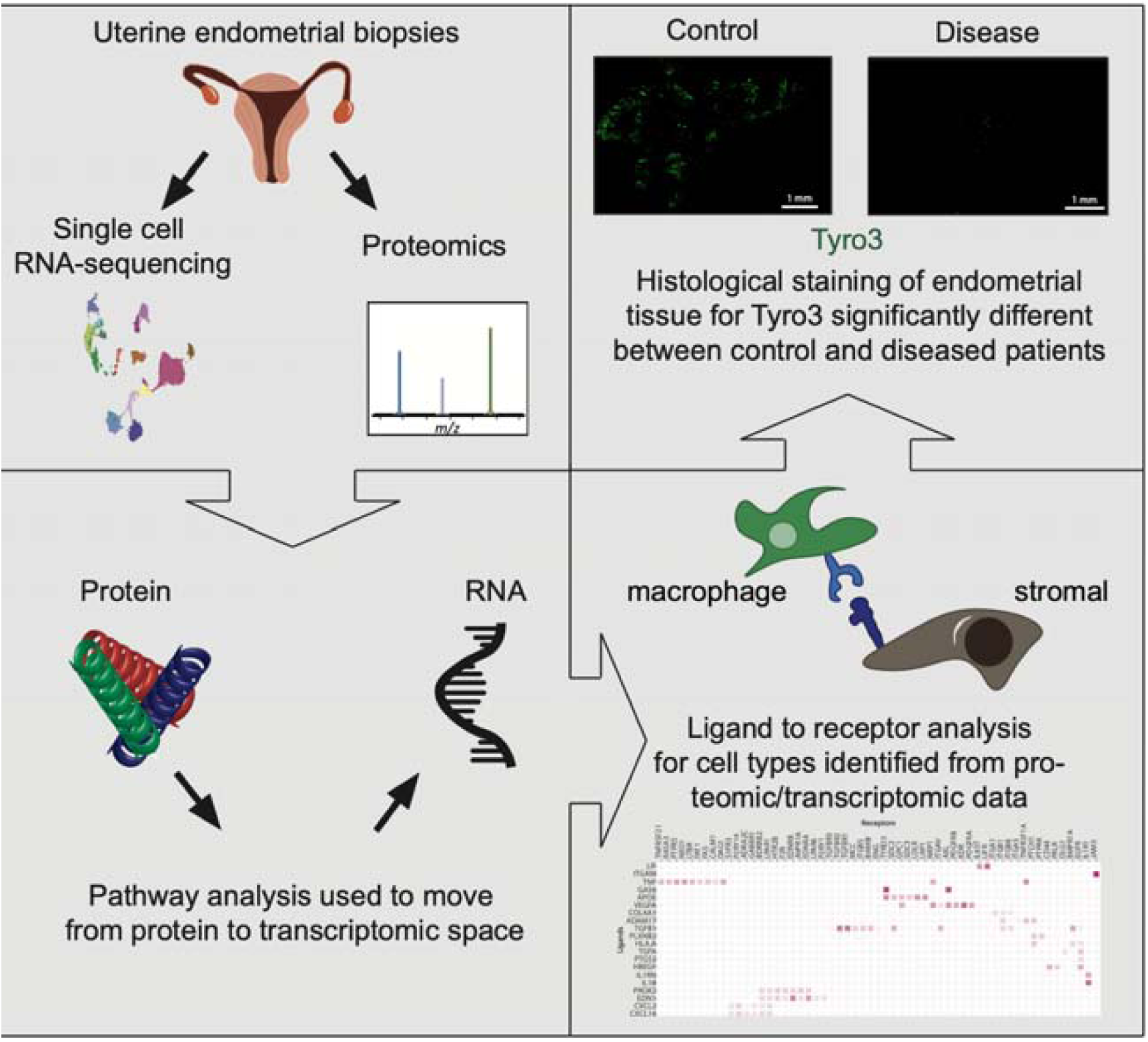

**Highlights:**

- Leverages proteomic data to interpret and direct single-cell RNA sequencing (scRNA- seq) analysis
- Demonstrates successful use of linear mixed effects models to attribute protein expression variance to either menstrual cycle phase or disease state
- Pathway analysis of disease state proteins guides identification of disease-relevant cell types present in scRNA-seq data, providing foundation for mining those data for receptor-ligand interactions of possible disease relevance
- A new potential non-hormonal target in endometriosis, TYRO3, emerges from confirming predictions of the receptor-ligand model with transcriptomic and immunohistochemistry analysis of an independent panel of endometrial biopsies

## 1. Introduction

Endometriosis is a debilitating inflammatory disease in which endometrial-like tissue grows outside of the uterus. An estimated 6-10% of all women and teens have endometriosis (Eskenazi and Warner, 1997), contributing to 35-50% of all pelvic pain and infertility (Meuleman et al., 2009), decreased quality of life, and imparting a societal economic burden of about $69 billion (Simoens et al., 2012). Despite these costs, medical limitations as well as social stigma delay diagnosis of endometriosis an average of over six years from the onset of symptoms (Nnoaham et al., 2011) with surgical intervention being the only definitive method of diagnosis. Adding to the difficulty in endometriosis diagnosis is the lack of clear medical distinction between lesion types, location, and symptoms as they relate to pain, infertility and other characteristics of the disorder (Zondervan et al., 2020). While there has been substantial progress made in the past several decades elucidating endometriosis pathophysiology, there is a clear need for better assessment and treatment options derived from a more nuanced understanding of disease mechanisms.

While there are many theories on the origin of endometriosis (Burney and Giudice, 2012), the most widely accepted is the century old theory of retrograde menstruation (Sampson, 1918). Sampson postulated that sloughed off eutopic endometrium, instead of exiting solely through the cervix during menstruation, also flows through the fallopian tubes into the peritoneal cavity leading to the development to endometriotic lesions (Sampson, 1918). Given that around 90% of women experience retrograde menstruation (Halme et al., 1984) – far more than the portion of women that develop endometriosis – this is likely only a small piece of the greater puzzle. There may be a hereditable component of the disease given that first-degree relatives of women with endometriosis have a six times greater risk of presentation compared to unaffected counterparts (Simpson et al., 1980) and genetic linkage studies have identified specific gene variants highly associated with disease (Tapmeier et al., 2021). Reports of molecular differences in the endometria of women with endometriosis compared to controls (Prašnikar et al., 2020) remain controversial (McKinnon et al., 2018), and such comparisons may require stratification to subpopulations of patients to be effective (Jørgensen, H.; Fedorcsak, P.; Isaacson, K.; Tevonian, E.; Xiao, A.; Beste, M.; Qvigstad, E.; Lauffenburger, D.; Griffith, 2022). One well characterized feature of endometriosis pathology is its response, or lack of response, to sex hormones; endometriosis is an estrogen dependent (Bulun, 2009), but progesterone resistant, disease (Burney et al., 2007). In practice, this makes studying the endometrium for possible mechanisms of endometriosis challenging because of the dynamic hormonal milieu throughout the 28-day idealized menstrual cycle.

About once a month, in the absence of pregnancy, the human uterus completes a menstrual cycle; a scarless wound healing processes in which the lining of the endometrium grows an average of 1-2 centimeters in thickness before being shed during menstruation. The cycle is typically broken into three phases involving the relatively stable stratum basalis and the outermost layer, the stratum functionalis, which is the region of the endometrium that undergoes the majority of the growth. The basalis and its resident cell populations support the fuctionalis as cells proliferate during the estrogen driven proliferative phase. The rise in progesterone following ovulation drives decidualization of the stromal fibroblasts, with an increase in extracellular matrix (ECM) production, during the secretory phase (Chan et al., 2004). Differences in cellular composition, RNA expression, and cellular response to stimulation in the menstrual effluent of women with endometriosis suggest that one, if not all, cycle phases may be aberrant with pathology (Warren et al., 2018), but nonlinear interactions throughout the menstrual cycle make it difficult to tease apart the variables.

The use of high-throughput biochemical assays has led to discoveries of potential biomarkers for endometriosis. Proteomic datasets have been used to identify at least 20 proteins that are aberrantly expressed in endometriosis patients (Ferrero, 2019). These proteins suggest dysregulation of the immune response as a major manifestation of endometriosis and hold promise for diagnostic development, however, these methods have not been implemented for clinical applications. Recently, transcriptomic analysis at single-cell resolution has opened up avenues for new and data rich methods of analysis that can classify cell populations, track cell states via lineage tracing, quantify cellular heterogeneity, and even infer cell-to-cell interactions to better understand disease pathology (Blencowe et al., 2019). However, with this wealth of information, it can be challenging to parse which cell types and transcriptomic profiles are significantly associated with disease pathogenesis, especially given the multiple variables (e.g., cycle phase of donor tissue, as we investigate here) in addition to the presence, or not, of endometriosis. Cycle phase is also difficult to control experimentally, as surgery is scheduled for availability of patient and surgeon, and attempts to pinpoint a specific cycle day can be complicated by variable cycle lengths in the patient, and cycles that do not conform to the idealized 28-day prototype.

To this end, the goal of this study was to deploy patient proteomic data, which functionally integrates transcriptional-level information, to direct the analysis and interpretation of single-cell transcriptomic data. For each protein, we constructed a set of linear mixed effect models (LMMs) to determine whether the expression was influenced by either the patient cycle phase or disease state. Next, we used pathway analysis on disease-relevant proteins to inform analysis of single-cell transcriptomic data, resulting in identification of cell types most relevant to the disease state. Then, guided by the known cell-cell communication networks in the endometrium, we performed directed cell-to-cell communication analysis with the cell types we identified by scRNA-seq analysis, seeking receptor to ligand interactions of possible relevance for disease phenotypes, and then confirmed findings on an independent sample cohort using immunohistochemistry. We found that the receptor tyrosine kinases Axl and Tyro3 and their respective ligands, Gas6 and Protein S, are altered in the endometriosis, identifying a further path of inquiry for potential clinical intervention.

## 2. Methods

### 2.1. Study approval

All participants provided informed consent in accordance with a protocol approved by the MassGeneralBrigham Human Research Committee and the Massachusetts Institute of Technology Committee on the Use of Humans as Experimental Subjects (Protocol number IRB-P001994). Endometrial pipelle biopsies were obtained from reproductive age women (N=17, ages 18-45) undergoing laparoscopic surgery for non-malignant gynecologic indications. Study enrollment was limited to pre-menopausal women and excluded patients with an irregular or ambiguous cycle history or a history of hormone use in the 3 months prior to surgery. A complete list of patients can be found in Supplemental Table 1.

### 2.2. Tissue Processing for scRNA-seq analysis

A single-cell suspension was generated from fresh endometrial Pipelle biopsies using enzymatic digestion and filter separation as previously described (Osteen et al., 1989). Briefly, the tissue was first minced and washed in 1X PBS to remove contaminating red blood cells (RBCs). Tissues were then incubated in a digest solution composed of collagenase type I (Worthington Biochem, Lakewood, NJ) solution containing DNAse I (SKU# 10104159001, MilliporeSigma, Burlington, MA) for 30 minutes in a 37°C water bath with frequent mechanical dissolution to separate the stromal cells from the epithelial glands. Isolated cells and fragments were then centrifuged and the pellet was resuspended in 5mL of Tryp-LE express (Cat# 12604013, Thermofisher, Waltham, MA) and DNAse I in a 37°C water bath for 15 minutes to generate a single cell suspension. Mechanical disruption of the cells was performed, and visual inspection of single cell populations was assessed using a microscope. Single-cell suspensions of endometrial cells were then either immediately processed for scRNA-seq or cryopreserved at -80°C in a cryopreservation (10% DMSO, 90% FBS) solution.

### 2.3. Tissue Preparation for Proteomics and Running the LC-MS/MS

Before digestion of the biopsy tissue for single cell analysis, a roughly 5 mm slice was isolated for proteomic analysis. Following collection, 50 μl of RapiGest (Product number 186001861; Waters Corporation, Milford, MA) reconstituted in 50 mM NH_4_HCO_3_ at a concentration of 2mg/ml was used to digest the biopsy sample for 24 hours at 37°C. Next, urea (Cat # 29700, Thermofisher, Waltham, MA) and dithiothreitol (Cat # R086, Thermofisher, Waltham, MA) were added to the sample to bring the mixture of a final concentration of 4 mM and 5 mM, respectively. This was left at 37°C for 24 hours in an incubator with a stir bar spinning at 1400 rpm. Finally, iodoacetamide (CAS 144-48-9, MilliporeSigma, Burlington, MA) was added to the sample, for a final concentration of 15 mM and kept in the dark, at 37°C, for 30 minutes. Samples were frozen at -80°C until all patient samples had been collected and processed.

Samples were lyophilized overnight and then reconstituted in NH_4_HCO_3_. A standard mixture was created by mixing equal parts from all of the patient samples into one tube. For each proteomics run, one aliquot of this standard mixture was used to normalize the sample readings between runs. Samples were digested with Trypsin Gold, mass spectrometry grade, from Promega (Madison, WI) using a ratio of 1 ug trypsin to 50 ug sample protein as quantified using a Pierce BCA Protein Assay Kit (Cat # 23225, Thermofisher, Waltham, MA). Samples were digested overnight at room temperature using a rotor for mixing and then placed in a speed vacuum to reduce sample volume. Dried down peptide samples were labeled with TMT10plex (Pierce, Rockford, IL, USA). After labeling, the samples were mixed and purified with peptide desalting spin columns (Pierce, Rockford, IL, USA). 1/5 of the sample was used for one LC/MS/MS analysis. The dried peptide mix was reconstituted in a solution of 2 % formic acid (FA) for MS analysis. Peptides were loaded with the autosampler directly onto a 50cm EASY-Spray C18 column (ES803a, Thermo Scientific). Peptides were eluted from the column using a Dionex Ultimate 3000 Nano LC system with a 5 min gradient from 1% buffer B to 10 % buffer B (100 % acetonitrile, 0.1 % formic acid), followed by a 44.8 min gradient to 25%, and a 9.2 min gradient to 36% B, followed by a 0.5 min gradient to 80% B, and held constant for 4.5 min. Finally, the gradient was changed from 80 % buffer B to 99 % buffer A (100% water, 0.1% formic acid) over 0.1 min, and then held constant at 99 % buffer A for 19.9 more minutes.

The application of a 2.2 kV distal voltage electrosprayed the eluting peptides directly into the Thermo Exploris480 mass spectrometer equipped with a FAIMS and an EASY-Spray source (Thermo Scientific). Mass spectrometer-scanning functions and HPLC gradients were controlled by the Xcalibur data system (Thermo Scientific). MS1 scans parameters were 120,000 resolution, scan range m/z 375-1600, AGC at 300%, IT at 50ms. MS2 scan parameters were either at 30,000 or 45,000 resolution, isolation width at 0.7, HCD collision energy at 36%, AGC target at 300% and IT set to auto. Cycle time for MS2 was 2sec for each MS1 scan.

All MS/MS samples were analyzed using Sequest (Thermo Fisher Scientific, San Jose, CA, USA; version IseNode in Proteome Discoverer 2.3.0.523). Sequest was set up to search uniprot_human_reviewed_032120.fasta (version March 21, 2020) assuming the digestion enzyme trypsin. Sequest was searched with a fragment ion mass tolerance of 0.020 Da and a parent ion tolerance of 10.0 PPM. Carbamidomethyl of cysteine and TMT6plex of lysine were specified in Sequest as fixed modifications. Oxidation of methionine and acetyl of the n-terminus were specified in Sequest as variable modifications.

Scaffold Q+ (version Scaffold_5.0.1, Proteome Software Inc., Portland, OR) was used to quantitate Label Based Quantitation peptide and protein identifications. Peptide identifications were accepted if they could be established at greater than 10.0% probability to achieve an FDR less than 1.0% by the Percolator posterior error probability calculation (Käll et al., 2008). Protein identifications were accepted if they could be established at greater than 99.0% probability to achieve an FDR less than 1.0% and contained at least 2 identified peptides. Protein probabilities were assigned by the Protein Prophet algorithm (Nesvizhskii et al., 2003). Proteins that contained similar peptides and could not be differentiated based on MS/MS analysis alone were grouped to satisfy the principles of parsimony. Channels were corrected by the matrix found in Supplemental Table 2 in all samples according to the algorithm described in i-Tracker (Shadforth et al., 2005). Normalization was performed iteratively (across samples and spectra) on intensities, as described in Statistical Analysis of Relative Labeled Mass Spectrometry Data from Complex Samples Using ANOVA (Oberg et al., 2008). Medians were used for averaging. Spectra data were log-transformed, pruned of those matched to multiple proteins and those missing a reference value, and weighted by an adaptive intensity weighting algorithm.

### 2.4. Linear Mixed Model for Proteomics Data

Linear mixed modeling (LMM) was used on the list of proteins discovered using isobaric tagging with LC/MS-MS. A total of 668 overlapping proteins were found and for each protein (Supplemental Table 3), a null model and an alternative model was built using the R package “lmer”. Protein expression was modeled by assuming that an individual protein response consisted of patient identity (Patient.ID) as a random variable and two vectors of fixed effect variables: *DiseaseState* - having endometriosis or not - or *cyclePhase* – the patient falling into either the proliferative or secretory phase. The two separate null models were built to assess the impact of cycle phase or disease state.

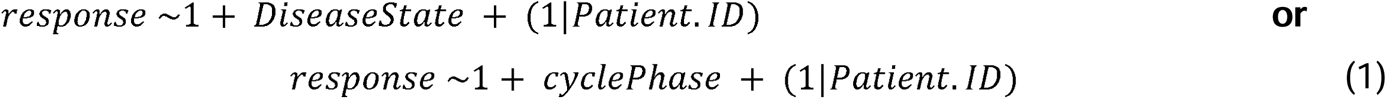

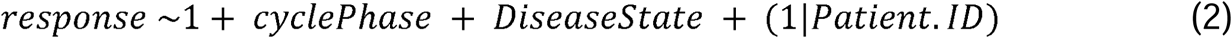

The each of the null models (1) were compared against the alternative model (2) using the likelihood ratio test (LRT).

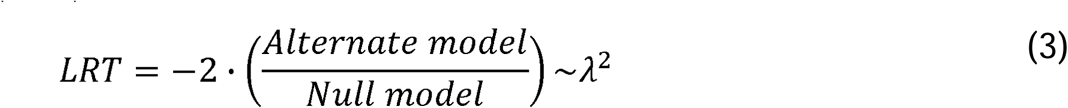

To determine the influence of cycle phase, the model was run once with disease state as the sole null model fixed variable and the alterative model including both cycle phase and disease state as fixed variables and, to determine the influence of disease state, the null model including cycle phase as the fixed variable. Patient identity was assumed to be a variable in all cases. As a way to generate hypothesis to test, significant proteins (P <0.05) were identified using the LRT. When using cycle phase as the null model fixed effect variable, 115 proteins were found to be significant. This was interpreted as those 115 proteins being significantly associated with disease state variance. Conversely, when disease state was the null model fixed effect variable, 43 proteins were identified as significant. Both proteins lists can be found in Supplemental Table 4 and scripts used for analysis are available for download here github.com/lauren-baugh in the LLM-Prot-Data repository. These identified protein lists were then used as inputs to look for significantly altered pathways using gProfiler (Raudvere et al., 2019) (Supplemental Table 5).

### 2.5. Generation of scRNA-seq data with Seq-Well S^3^

Seq-Well *S*^*3*^ was performed as described previously.(Gierahn et al., 2017; Goods et al., 2021; Hughes et al., 2020) Briefly, at least 15,000 cells were loaded onto each array preloaded with barcoded mRNA capture beads (ChemGenes). Arrays were washed with protein-free RPMI media, then sealed with plasma-treated polycarbonate membranes. Arrays were incubated at 37°C for 30 minutes to allow membranes to seal, then transferred through a series of buffer exchanges to allow for cell lysis, transcript hybridization, bead washing, and bead recovery from arrays after membrane removal. Reverse transcription was performed with Maxima H Minus Reverse Transcriptase (ThermoFisher) and excess primers were removed using an Exonuclease I digestion (New England Biolabs). Second strand synthesis was performed and whole transcriptome amplification (WTA) by PCR was performed using KAPA Hifi PCR Mastermix (Kapa Biosystems). WTA product was purified using Agencourt Ampure beads (Beckman Coulter) and 3’ digital gene expression (DGE) sequencing libraries were prepared using Nextera XT (Illumina). Libraries were sequenced using custom primers on a Novaseq S4.

### 2.6. Computational analyses of scRNA-seq data

#### Alignment and cell identification

Raw sequencing data was demultiplexed and aligned to the Hg19 genome using publicly available scripts on Terra (scCloud/dropseq_workflow, version 11, https://app.terra.bio/). Raw sequencing data will be made available for download concurrently with publication on the Gene Expression Omnibus database. The resulting UMI-collapsed digital gene expression matrices were used as input to Seurat (v3) for further analyses in R (v3.6). All data were normalized, the top 2,000 variable genes identified. Initial clustering was performed and cell types were assigned on the basis of marker genes identified using Seurat’s Wilcoxon rank-sum test and comparison to those in the existing literature (Supplemental Table 6). After initial analyses, doublets were removed using DoubletFinder following default settings (McGinnis et al., 2019), cells were re-clustered, and cell types were finalized based on final cluster markers (Supplemental Table 6). We also iteratively sub-clustered immune cells in order to refine cell type identification on the basis of a new set of marker genes that was distinct from those driven by epithelial, stromal, and immune cell global differences. All scripts to reproduce analyses are available for download (https://github.com/bagoods).

#### MAST modeling

In order to identify differentially expressed genes in the major cell populations that were driven by either cycle stage or disease, we used MAST modeling (Finak et al., 2015), as this approach performs well for complex technical variables with fast runtimes (Luecken and Theis, 2019). We performed differential expression on epithelial cells and stromal cells separately with the following model: zlm(∼CycleStage + DiseaseStatus) (Supplemental Table 7 and 8). Next, for each cell type we filtered for significant genes on the basis of >0.3 fractional expression and padj < 0.05 for either CycleStage (secretory vs proliferative) or DiseaseStatus (endometriosis vs healthy). MAST also produces regression coefficients that indicate the direction and relative magnitude of fits of the hurdle model for each gene. To simultaneously explore the relationship on a gene-gene level, we used ggpubr() to create scatter plots of these coefficients. All scripts to reproduce analyses are available for download (https://github.com/bagoods).

#### NicheNet modeling

scRNA-seq expression data was sub-set by each patient then saved as separate R objects. NicheNet (Browaeys et al., 2020) analyses were performed using standard workflows on patients 259, 260, 270 and 271 since these patients had enough macrophages detected to run this type of analysis. NicheNet analysis was run in two modes for each patient separately. First, we wanted to determine which signals from macrophages were impacting variable gene expression in either epithelial cells or stromal cells. NicheNet’s ligand-target prior model, ligand-receptor network, and weighted integrated networks were imported from a publicly available database (https://zenodo.org). The gene set of interest was defined by normalizing the expressed genes by the assigned receiver cell type then finding the most variable features. For each patient, variable genes were identified with Seurat within the largest cluster of either stromal or epithelial cells. After defining a set of potential ligands using the imported network data, they were ranked by the presence of genes downstream of their target receptors in the receiver epithelial or stromal cells. These top-ranking ligands were then analyzed by inferring receptors and target genes to produce a heatmap to show the regulatory potential between prioritized ligands and their predicted target genes/receptors. All scripts to reproduce these analyses are available for download (https://github.com/bagoods).

### 2.7. Tissue Sectioning, Staining, and Quantification

#### Histological tissue sectioning

Histological slices of the patient biopsy samples were fixed with 4% paraformaldehyde upon retrieval from Newton-Wellesley Hospital. After fixation, samples were dehydrated and cleared with xylenes before being paraffin embedded. Slides were produced by slicing the tissue block into sections using a microtome. To prepare for histological staining, the slides were deparaffinized using xylene and a reducing series of graded alcohol.

#### Immunological staining

Slides were blocked for 4 hours at room temperature using a solution of 3% bovine serum albumin (BSA; CAS 9048-46-8, MilliporeSigma, Burlington, MA) and stained using the capture antibody from the Tyro3/Dtk DuoSet Elisa kit (Cat # DY859, R&D Systems, Minneapolis, MN) at a concentration of 25 μg/mL and CD45 antibody (Cat # MA5-17687, Thermofisher, Waltham, MA) using the recommended 1:500 dilution for 48 hours at 4°C. Secondary antibodies anti-mouse Alexa Fluor 488 and anti-rat Alexa Fluor 546 (Cat #s A-21202 and A-11081, Thermofisher, Waltham, MA) were used at a concentration of 10 μg/mL along with 4’,6-Diamidino-2-Phenylindole, Dihydrochloride (DAPI; Cat # D1306, Thermofisher, Waltham, MA) to stain for nuclei. Slides were washed with 1X phosphate buffered saline (PBS; Cat # 10010023, Thermofisher, Waltham, MA) and stained for 3 hours at room temperature. Slides were then washed again with 1X PBS and imaged using a Keyence BZ-X700 series microscope using a 10x objective.

#### Image quantification

Images were quantified using CellProfiler(McQuin et al., 2018) to identify the number of CD45+ cells and the total area that stained positive for Tyro3. Brightfield images were used to determine the area of each tissue sample and this measurement was used to normalized CD45 and Tyro3 expression between patients. Unpaired student t-tests were used to evaluate significance of CD45 expression and Tyro3 expression between control and diseased patient populations.

## 3. Results

### Generation of scRNA-seq and proteomics data in healthy and endometriosis patients

In order to develop a more complete proteomic and transcriptomic picture of the healthy and diseased uterus, we established a workflow to profile human endometrial samples. Endometrial biopsy samples were collected from patients undergoing hysterectomy. Prior to uterine tissue removal, pipelle biopsies are taken, primarily from the fundus region of the uterus. These biopsies were then digested as described above to prepare samples for scRNA-seq and proteomic (Figure 1A) analyses. Patients without a history or diagnosis of endometriosis were used as control subjects while diseased patients presented with symptoms of endometriosis with an official diagnosis made during or after surgery. In total, we profiled 11 total patients using scRNA-seq (n=6) and proteomics (n=8). For our proteomic data, we first performed principal component analysis across 668 proteins. Displaying this data along the first and second dimensions revealed little global separation on the basis of cycle phase and of disease state of the patient samples (Figure 1B). There is minor separation of patient disease state along principal component (PC) 1, driven by differences in extracellular matrix composition since COL3A1, COL6A1, and RACK1 were in the top 10 of PC1 contributors.

**Figure 1.**
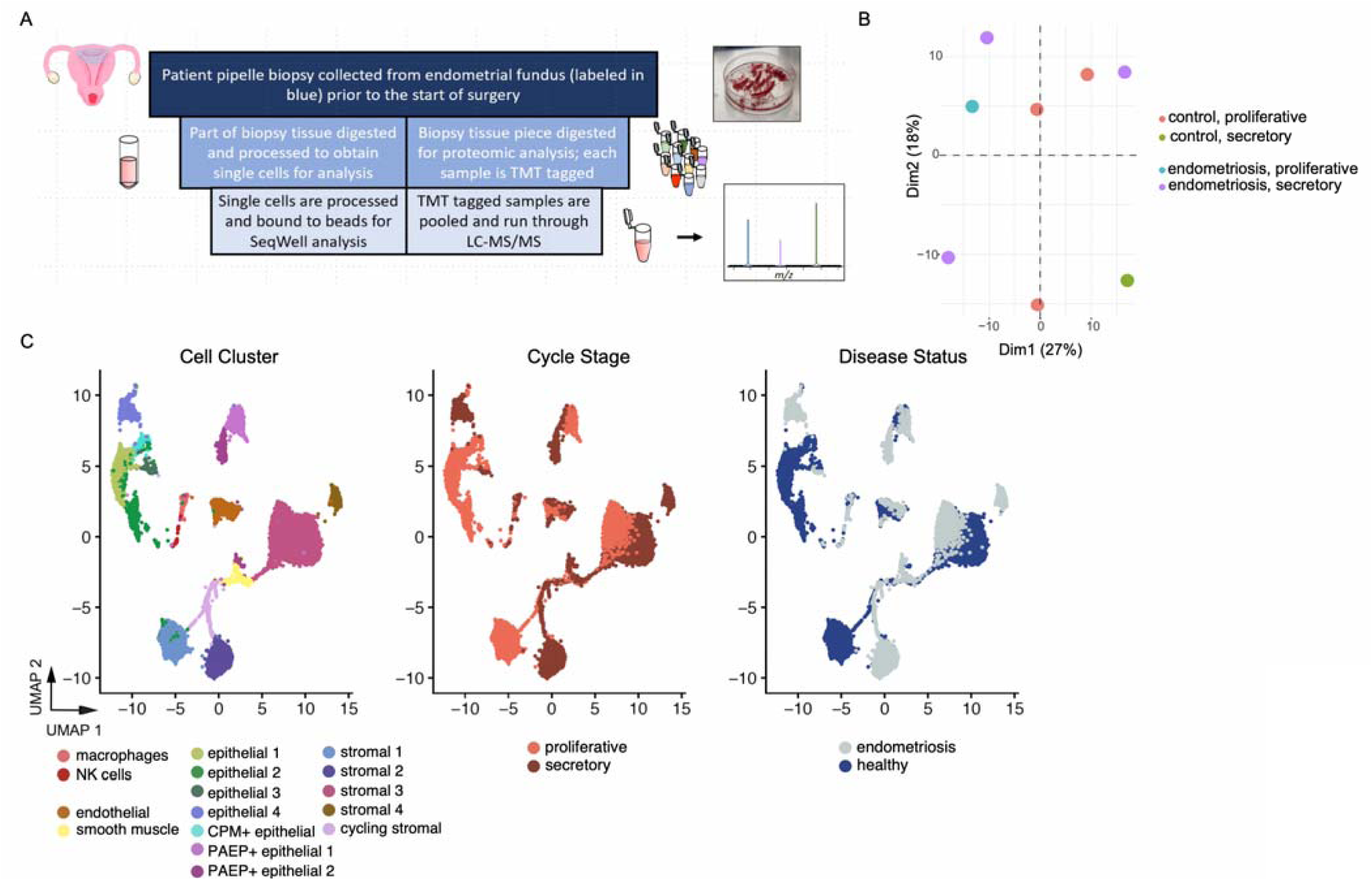
Sample collection workflow and study overview. **A**. Sample collection workflow. Endometrial biopsies are collected via pipelle from women undergoing uterine surgery. The sample is split, with part used to isolate single cells for scRNA-seq analysis using Seq-Well, and the remainder digested and analyzed for proteomics using LC-MS/MS. **B**. A PCA plot of the proteomics samples show poor clustering based on disease state with cycle phase as a confounding factor (n = 8 donors, Supplementary Table 1). C. ScRNA-seq data (n = 6 donors, 6,659 cells) shown as a UMAP projection colored by cell identities (left), cycle stage (middle) or disease status (right).

We generated high quality scRNA-seq data on a total of six patients (3 control, 3 endometriosis) and 6,659 single cells, and identified major epithelial, stromal, and immune populations across all six donors (Supplemental Figure 1). Dimensionality reduction of the scRNA-seq data using PCA followed by uniform manifold approximation and projection (UMAP) on dimensions 1 and 2 highlights populations of cells that cluster strongly by both cycle phase (center, Figure 1C) and disease state (left, Figure 1C). Taken together, our data suggest that both cycle and disease state drive global variations in both data types, and suggests that analytical approaches that leverage both these variables are essential for deeper analyses.

### Identification of cycle phase and disease associated proteins

Initial unsupervised clustering methods for both the proteomic and scRNA-seq data highlighted the difficulties of having both menstrual cycle phase and disease state as variables of interest. We therefore used linear mixed effect modeling (LMM) in order to tease apart the effects of these two biological variables in our proteomics results. Using a LMM, we attempt to capture the majority of variance in the system using either fixed or random variables and then by creating a null model to capture for comparison we isolate the variance attributed to the variable of interest. This allows us to assess individual factors while avoiding the pitfalls of trying to understand the contribution of each source of system variance. The likelihood ratio test was used between a null model and an alternative model for each protein as described (see Methods). The model was run twice to generate a list of proteins that showed significant association with cycle phase or disease state. Of the 669 proteins identified from the biopsy tissue, 115 were significantly affected by disease state and 43 were significantly affected by cycle phase, with 28 overlapping proteins. Volcano plots of the identified protein lists show more proteins are upregulated in disease state compared to those that are downregulated (or proteins upregulated in control patients) and more proteins are significantly upregulated in the secretory phase as opposed to the proliferative phase (Figure 2A). PCA of the refined set of proteins from the patient proteomic data separates groups based on disease state (Figure 2B, left) and cycle phase (Figure 2B, right). This suggests that the use of LMM was successful in deconvolving the impacts of cycle phase and disease state, revealing a set of proteins associated with each to investigate further. While numerous protein differences among samples occur as a result of the natural menstrual cycle, an independent set of proteins are associated with the disease state, and a subset of proteins is significantly affected by both cycle phase and disease state.

**Figure 2:**
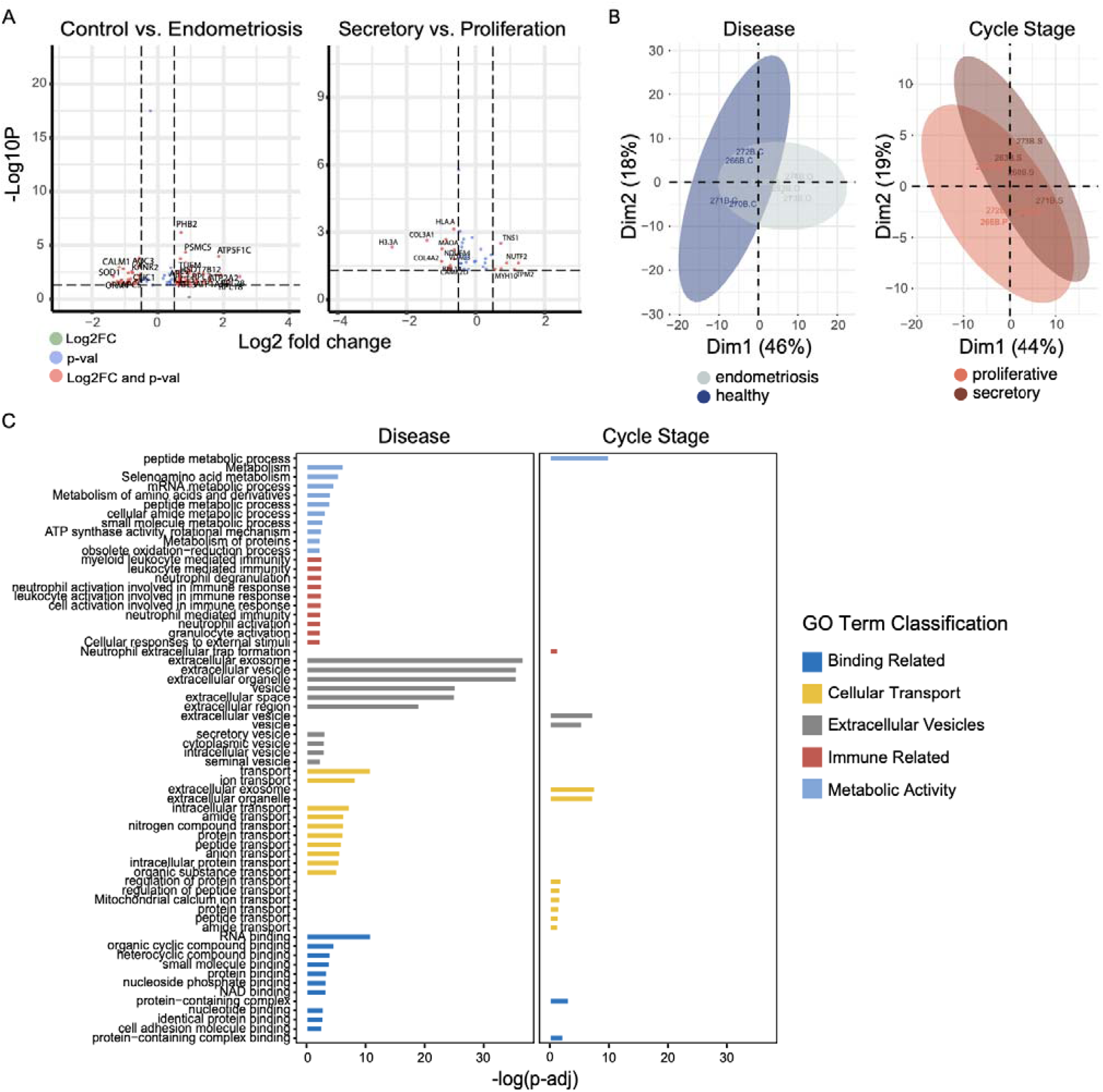
Linear mixed effect modeling (LMM) to identify variable proteins across cycle stage or disease status. **A**. Volcano plots of the proteins identified in the LMM using either disease state or cycle phase only in the null model. Green points are above the log_2_ fold change cut, blue points are above the -log_10_ of the p-value cut off and red points are above both the - log_10_ of the p-value and log_2_ fold change cut offs. **B**. LMM is used to identify proteins that are variable either in disease state or with cycle phase. In the LMM for each protein, the null model excludes the variable of interest (disease or cycle phase) and is compared against the full model to identify proteins with a significant impact on the alternative model variable. PCA plots using the proteins identified in the two different models show better separation of both groups. **C**. A heatmap of functional enrichment results on protein lists identified as significant in disease or cycle phase.

In order to determine which signaling pathways might be influenced by either cycle stage or disease status, we performed pathway analysis on the LMM protein lists using the software gProfiler (Raudvere et al., 2019). We identified hundreds of pathways that were altered across both protein lists (Supplemental Table 4). To better understand the broader implications of these results, we classified these pathways based on each GO terms description: immune response, extracellular vesicles, cellular transport, cellular binding, and cellular metabolism. We show the top ten scoring pathways in each broad category (Figure 2C). These data suggest that pathways significant to cycle phase skew significantly towards cellular transport pathways while disease-related pathways are notably higher in the immune response, extracellular vesicles, cellular binding, and cellular metabolism categories (Figure 2C). Taken together, our analyses suggest that we are able to effectively tease apart differential proteins and associated biological functions that differ as a function of cycle stage or disease status using this LMM approach.

### Multiple cell types are involved in expression of proteomics-identified genes

We next sought to leverage our scRNA-seq data set to better understand our proteomics results by determining which cell types were expressing cycle or disease-defined transcripts. We found that cycle-associated genes, including TPM2, TPM1, COL4A2, and TNS1, were predominantly expressed in smooth muscle cells (Figure 3A). We also found that immune related genes, such as HLA-A, were expressed in macrophages and NK cells. For those proteins that were identified as uniquely associated with disease, we found that many different cell types expressed the corresponding genes (Figure 3B). These included expression of IDH2 across several different clusters of fibroblasts and epithelial cells, high expression of SOD2 in epithelial cells, and high expression of LCP1 in macrophages and NK cells. We additionally use a MAST (Finak et al., 2015) modeling approach to identify genes associated with both cycle and disease in epithelial cells and stromal cells since these were the largest cell populations in our dataset (Figure 3C, Supplemental Table 7 and 8). We identified many genes that were significantly differentially expressed across both disease and cycle, including many that were found in our proteomi data. Comparing our disease-associated proteins with the corresponding lists in our scRNA-seq data, we found there were 71 common elements in epithelial cells and 69 in stromal cells. Similarly, for cycle-associated elements, we found 15 common for epithelial cells and 20 common for stromal cells. Overall, this suggest that many different cell types are contributing to disease and cycle associated alterations in the uterus, and by combining both scRNA-seq and proteomics, we can identify high-confidence lists of genes of interest.

**Figure 3:**
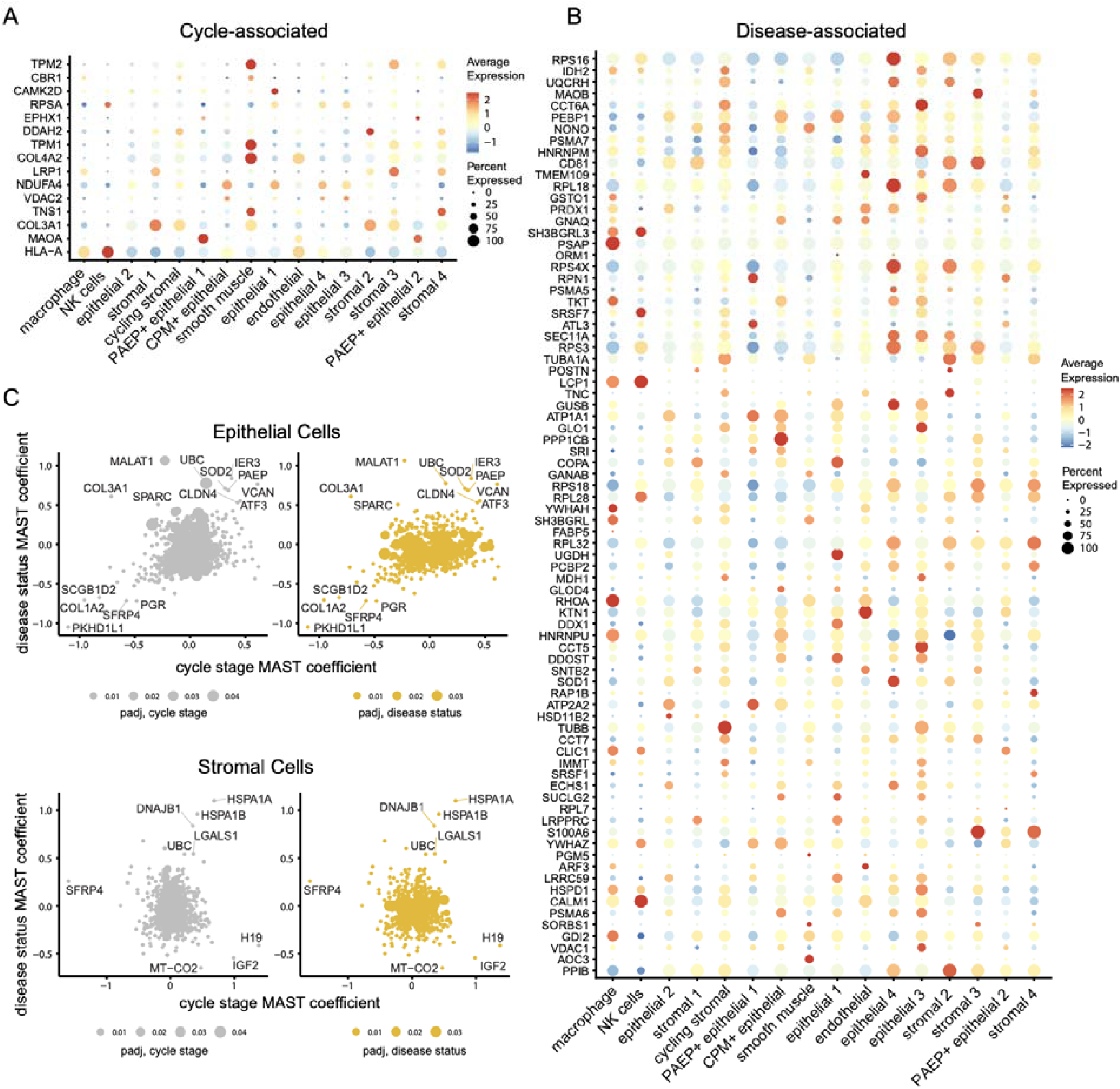
Expression of cycle-associated or disease-associated genes in scRNA-seq data. **A**. Dotplots of genes uniquely associated with cycle stage as identified in Figure 2 as shown across all cell identities. **B**. Dotplots of genes uniquely associated with disease status as identified in Figure 2 shown across cell identities. **C**. Scatter plots of MAST modeling coefficients where each dot is a gene and scaled according to the magnitude of the adjusted p value.

In order to look at these lists more wholistically, we plotted the MAST regression coefficients for cycle stage and disease for either epithelial cells or stromal cells (Figure 3C). These plots allow us to identify genes that significantly contribute to disease or cycle and the magnitude of contributions. For example, we see that for epithelial cells, UBC, SOD2, VCAN, and PAEP were found to be significantly positively associated with cycle stage and disease, while COL1A2 and PGR were both negatively associated. We additionally can see that there are fewer genes associated positively with disease and cycle stage in stromal cells, but they include several heat shock protein genes and LGALS1. The long non-coding RNA H19 was found to be associated positively with cycle stage in stromal cells. Taken together, these plots allow us to further identify genes associated with cycle stage and disease in the uterus and serve as a roadmap for deconvolving the complex interplay between genes associated significantly with both.

### Enrichment of proteomics-identified pathways across scRNA-seq data

Next, we wanted to determine which cell types in our scRNA-seq data were enriched for top-scoring pathways identified in our proteomics data. To do this, we created module scores for proteomics-identified pathways and performed PCA on the resulting scores (Figure 4A). We found that we could separate cell types along PC1 or PC2 based on each cell type’s high-scoring pathways. We found that macrophages separated from the rest of the cells along PC2, where the top pathways driving this separation were immune in nature, including myeloid leukocyte mediated immunity (Figure 4B), neutrophil activation, and leukocyte mediated immunity. We additionally found that there was a spectrum of epithelial and stromal cells, with the epithelial 4 cluster and PAEP+ epithelial 2 cluster separating along PC1. This was predominantly driven by selanoamino acid metabolism and regulation of protein transport (Figure 4B). This suggests that there might be different functions of these epithelial and stromal subsets, and in light of other published reports (Tan et al., 2021; Wang et al., 2020), suggests that proteomics coupled with scRNA-seq can help to refine these phenotypes. Taken together, our data suggest that this approach can provide useful context for interpreting long lists of pathway enrichment results from proteomics data and also identify subsets of cells, like stromal, epithelial, and macrophages that drive these results.

**Figure 4.**
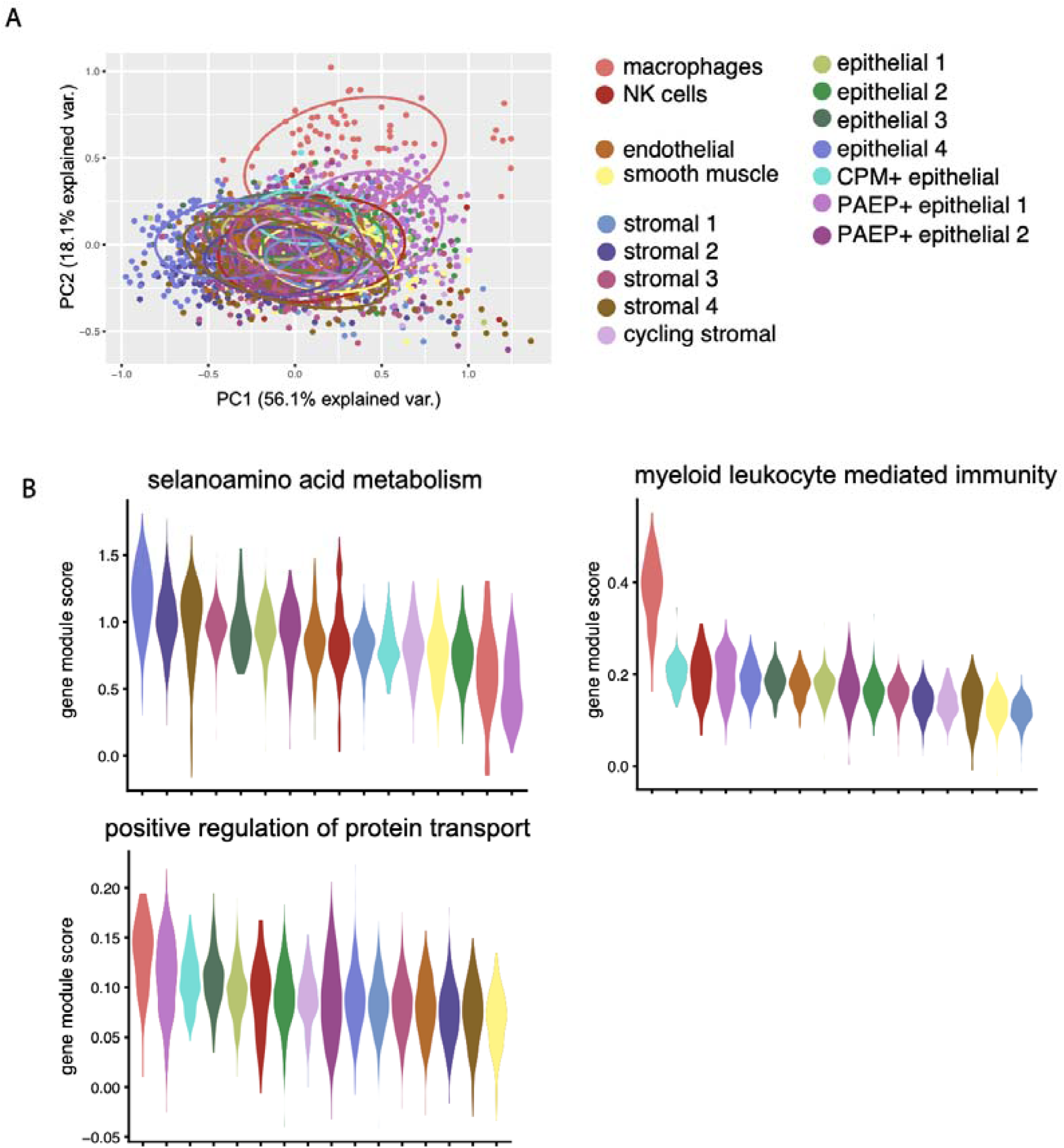
Module scoring identifies cell types that may drive pathway enrichments from proteomics data. **A**. PCA plot of module scores calculated for each top scoring pathway from Figure 2D. PCA plot is colored by cell type. **B**. Violin plots of the top variable loading pathways along PC1 and PC2.

### Mining interactions between stromal cells or epithelial cells and macrophages suggests Axl-mediated signaling is altered across disease status

Given that our proteomic and scRNA-seq analyses suggested that macrophages, stromal cells, and epithelial cells have different roles in cycle and disease-related uterine biology, we wanted to determine how cell-cell interactions might influence these processes. To do this, we used NicheNet, (Browaeys et al., 2020) a cell-to-cell ligand-to-receptor interaction inference approach, to identify receptors and ligands involved in specific interactions identified strongly with disease state (see Methods). We performed this on a patient-by-patient basis in order to more accurately identify interactions between macrophages and stromal or epithelial cells in a subset of patients that had enough macrophages present in the dataset (see Methods). We identified several major receptor and ligand interactions that may be altered in the context of disease, health, and as a function of patient. For interactions between macrophages and epithelial cells, we found several receptor-ligand pairs with high interaction potentials that were conserved across all patients, including IL1B-IL1R, and several that were distinct across patients, such as APP interactions with various receptors (Supplemental Figure 2). Interestingly, for macrophage and stromal cell interactions (Figure 5), we found that Axl-GAS6 receptor-ligand pair scored highly across all patients. We previously found in our MAST modeling results that Axl was differentially expressed in both cycle and disease conditions in stromal cells as well (Supplemental Table 8). Axl is part of group of tyrosine kinase transmembrane receptors which also includes MERTK and Tyro3, collectively known as “TAM” receptors. They are known for their roles in efferocytosis and immune checkpoint blockades (Lemke, 2019; Lemke and Rothlin, 2008; Vago et al., 2021). Interestingly, we also see Gas6-Tyro3 binding between macrophages (Gas6) and stromal cells (Tyro3) as important cell to cell interactions, though this ligand receptor pair is less well studied compared to Axl. Taken together, this data suggests that macrophage interactions with stromal cells, as mediated by Axl and Tyro3 signaling may play a key role in uterine cycle and disease.

**Figure 5.**
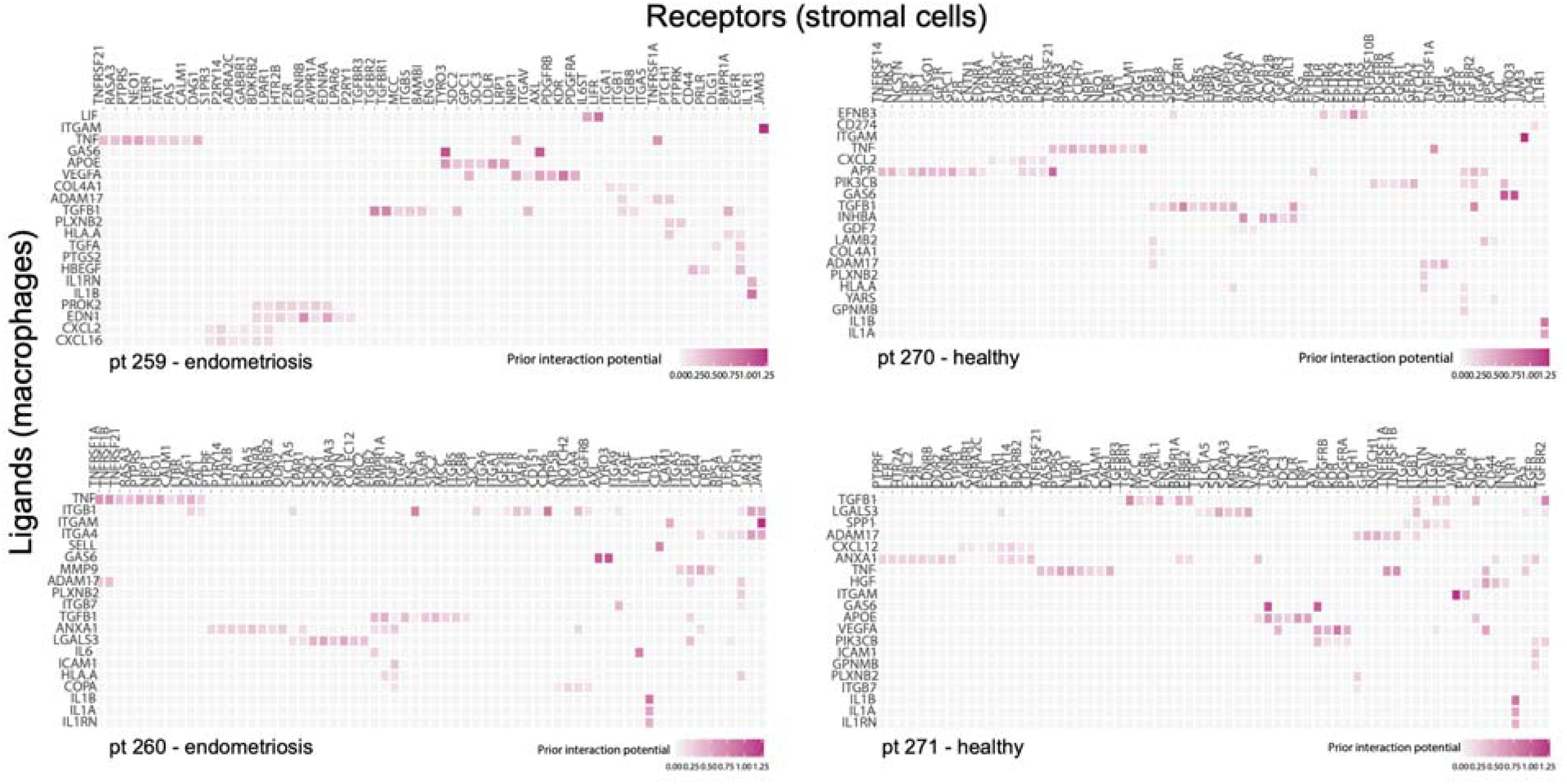
NicheNet modeling results suggest key receptor and ligand pairs that mediate immune-stromal interactions. For each patient, top predicted ligands and receptors are shown for macrophages interacting with stromal cells.

### Axl and Tyro3 mediated signaling in the healthy and diseased uterus

We next wanted to further investigate the role of Axl, Gas6 and Tyro3-mediated signaling in the healthy and diseased uterus. Further analysis of Axl (data not shown), Gas6 and Tyro3 expression in our scRNA-seq data of all patients shows that Tyro3 is downregulated in patients with disease while Axl and Gas6 do not follow this trend (Figure 6A, B). Following up on Tyro3, and one of its primary ligands found to be expressed in macrophages (from Figure 4) Gas6, expression of Gas6 does not seem to have a notable difference between disease states in patients with a measurable macrophage population (data not shown), while Tyro3 expression in stromal cells, and a lesser degree epithelial cells, is downregulated significantly in diseased patients (Figure 6A and B). To confirm these results and better delineate the role of Tyro3, we used immunohistochemistry on slides of patient endometrial biopsy tissue (representative image in Figure 6C) to stain for Tyro3. Tyro3 staining was found throughout all samples in controls, but was rare in endometriosis patient specimens (Figure 6D). We used CD45 to quantify immune cell populations between patient groups, capturing macrophages and other immune cell types. CD45 expression is comparable to values previously reported (Jones et al., 1996; Vallvé-Juanico et al., 2019) and does not differ between populations as shown in Figure 6E and quantified in Figure 6F. This, combined with our single-cell transcriptomic results, suggests that dysregulation of Tyro3 resides in endometrial stromal cells; however, more detailed analysis is required to confirm this finding.

**Figure 6.**
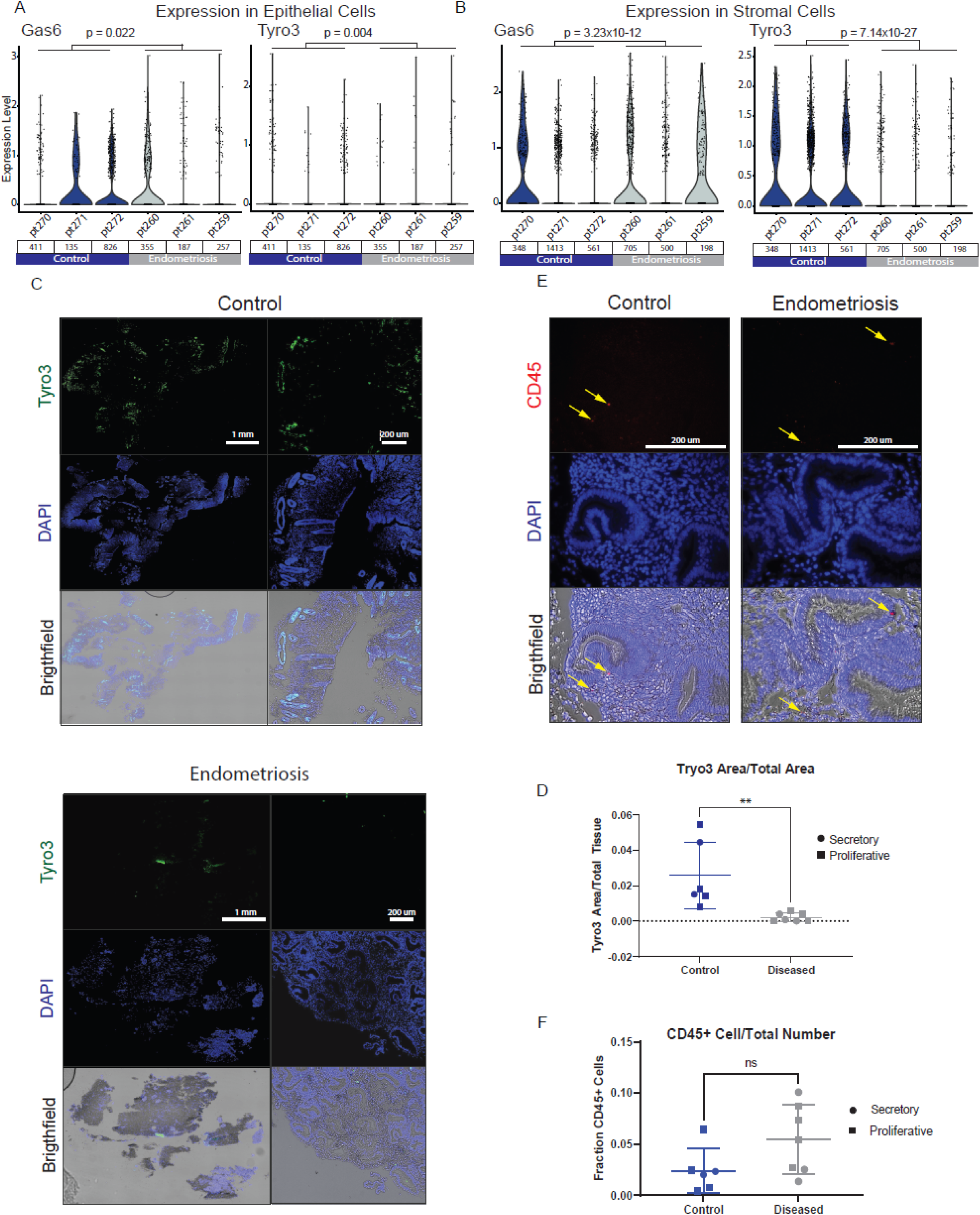
Expression of CD45 and Tyro3 in endometrial tissue samples show no difference in immune cell populations, but Tyro3 is downregulated in disease patients. **A**. Gas6 and Tyro3 expression across all epithelial cells. P values were derived from MAST modeling (Supplemental Table 7). **B**. Gas6 and Tyro3 expression across all stromal cells. P values were derived from MAST modeling (Supplemental Table 8). **C**. Representative images of histological staining for Tyro3, DAPI, and an overlay with brightfield (bottom) in patient endometrial biopsy samples are quantified in **D** by comparing the area of Tyro3 staining normalized by the entire tissue area. There is a significant decrease in Tyro3 expression in diseased patients. **E**. Histological staining for CD45 (some cells marked with yellow arrows), quantified in (**F**), show no significant difference in CD45 expression

## 4. Discussion

Although endometriosis was first recognized clinically by Sampson over 100 years ago (Sampson, 1918), there has been little headway in developing robust diagnostic capabilities or treatment options beyond hormonal regulation, and surgery. Part of the difficulty in identifying mechanism and biomarkers of the disease stems from the dynamic hormonal environment of the human uterus along with a complex and intricate tissue and immune cell network. New methods for detailed characterizations of tissues, like scRNA-seq, can provide a wealth of information about cell states and their functions in tissues (Ordovas-Montanes et al., 2020). Indeed, previous scRNA-seq studies have characterized cells over the course of the menstrual cycle and revealed genes and cells associated with the window of implantation(Wang et al., 2020), as well as guided endometrial organoid development via detailed understanding of cellular differentiation trajectories (Garcia-Alonso et al., 2021). While scRNA-seq data provides a wealth of information that can help tease apart intricacies associated with disease, the task of deciphering out relevant biological implications can be daunting. By leveraging more diagnostically-friendly proteomic data to direct the search of cell types and cellular mechanisms associated with endometriosis, we have identified new potential avenues for disease diagnosis and treatment.

The use of LMM on our proteomic data identified lists of proteins that were significantly associated with either cycle phase or disease. These selected lists were then able to separate out the patients based on the variable used in the modeling selection using the unsupervised, dimensionality reducing PCA (Figure 2B). As further validation of the accuracy of the LMM model, proteins found to vary with cycle phase were compared to other proteomic studies of the menstrual cycle. While there is direct overlap of proteins like Collagen IV, there is also overlap in identifying pathways associated with the menstrual cycle such as cellular transport.(Rai et al., 2010) Since direct protein to RNA comparison can be limited in accuracy,(Vogel and Marcotte, 2012) we sought to identify differentially expressed genes that were also found in our proteomics LMM results. We additionally used pathway analysis to bridge this gap, and found that we could nominate cell types that were potentially driving enrichment results. This further allowed us to narrow down on cell types for further investigation, focusing on macrophages, stromal cells, and epithelial cells.

Many of the disease relevant proteins, when projected to the single-cell transcriptomic data were expressed in macrophages, epithelial cells, and stromal cells over varying phases of the menstrual cycle (Figure 3B). Previous studies have also identified these cells types to be important in endometriosis as both the immune system and endogenous endometrium have been implicated in the disease. It was recently discovered that macrophages from people with endometriosis have increased expression of SIRP-α suggesting that these macrophages may have an impaired ability to phagocytosis cells.(Xie et al., 2019) Given the importance of macrophages in the clearance of shed menstrual effluent, the inability to properly breakdown the stromal and epithelial cells present in these tissue effluents could exacerbate endometriotic lesions. In addition to associated aberrations in the immune system in women with endometriosis,(Ahn et al., 2015; Symons et al., 2018) there is also research implicating both epithelial cells and stromal cells. This includes increased stem-cell like features(Hapangama et al., 2019) and higher expression of stimulator of interferon gene (STING)(Qu et al., 2020) in epithelial cells in women with endometriosis. While stromal cells have not been shown to have stomatic mutations related to endometriosis (Moore et al., 2020), they may have epigenetic abnormalities such as a decrease in progesterone receptor expression causing progesterone resistance,(Attia et al., 2000) though many of these stromal cell factors are found to be aberrant in the lesions themselves rather than the endometrium of patients.(Bulun et al., 2019) Additionally, our MAST modeling approach suggests that there are many genes that contribute to both cycle and disease-related attributes (Figure 3D). Here, we have provided an approach and combined dataset that has nominated many more genes that require further study and validation.

With the focus on three identified cell types and the pathway enrichment score from the proteomics data suggesting that cellular crosstalk is upregulated in the endometrium of patients with endometriosis, we analyzed ligand receptor interactions of macrophages to epithelial cells and stromal cells (Figure 4). For the four patients analyzed, the consistently strongest ligand receptor pairs were found in the macrophage to stromal cells interaction with macrophages expressing growth arrest – specific 6 (Gas6) and stromal cells expressing the receptor tyrosine kinases Axl and Tyro3. These two receptors are part of a group of transmembrane receptors which, together with MERTK, make up the TAM (Tyro3, Axl, and MERTK) receptors that are known for their roles as immune check point inhibitors and in efferocytosis.(Lemke and Rothlin, 2008) The TAM receptors are known to bind to both Gas6 and Protein S,(Nagata et al., 1996) though Gas6 has the highest affinity for Axl while Protein S has the highest affinity to Tyro3.(van der Meer et al., 2014; Nakamura et al., 1998) Further inspection of the transcriptomic data did not show a notable trend with Gas6 or Axl expression, but expression levels for Tyro3 were decreased in patients with endometriosis (Figure 6) and Protein S followed a similar transcriptomic profile to Tyro3 (Supplemental Figure 3). An increase in TAM receptor expression with inflammation as an anti-inflammatory response(Lemke, 2013) and increased TAM expression would presumably help with phagocytosis in the endometrium, however, both phagocytosis(Weng et al., 2020) and inflammation(Lin et al., 2018) have been documented as abnormal in patients with endometriosis. Additionally, TAM^-/-^ mice have been shown to develop autoimmune disorders,(Li et al., 2013) with this response dependent on the number of receptors that have been knocked out.(Lu and Lemke, 2001) Endometriosis also shares many characteristics with autoimmune disorders and patients with endometriosis have higher instance of autoimmune disease.(Shigesi et al., 2019) Our results indicate that the measured decrease in Tyro3 expression in patients with endometriosis compared to controls could be used as a biomarker – with lack of Tyro3 indicating an increased risk of endometriosis. Furthermore, this key inference highlights the TAM receptors as potentials treatment avenues for patients suffering from or prone to endometriosis as an increase in TAM receptor signaling in diseased patients may promote a more effective immune response to menstrual effluent.

Follow up studies to explore TAM receptors and their potential role in endometriosis could include additional patient data sets to gain more granular insights in cell expression across the phases of the menstrual cycle, as our limited single cell transcriptomic data suggests that Tyro3 expression may be cycle phase dependent. The striking difference in observed Tyro3 immunohistochemistry outcomes between control and endometriosis patients – where the greatest observed staining values in endometriosis patients are still below the lowest values in controls – could arise from a combination of transcription and post-translational events such as proteolytically-mediated shedding. Tyro3 undergoes enhanced shedding, producing prodigious amounts of soluble receptor, in rheumatoid arthritis (Vullings et al., 2020). These mechanisms could be further parsed in 3D co-cultures, building on advances in 3D tissue engineering models that incorporate the relevant cell types in synthetic ECMs (Below et al., 2022; Gnecco et al., 2021).

In summary, we used linear mixed effect modeling to decipher the effects of menstrual cycle phase and disease state and used these identified proteins to discover pathways specific to endometriosis and cycle phase. This pathway analysis then guided single-cell transcriptomic analysis to parse dysregulated cell-to-cell interactions between macrophages and stromal cells. The TAM receptor Tyro3 was shown to be downregulated in patients with endometriosis, at both the transcriptomic and protein level, paving the way for further analysis on the use of Tyro3 as both a diagnostic and treatment pathway.

## Data Availability

All data produced in the present study are available upon reasonable request to the authors

## References

Ahn, S.H., Monsanto, S.P., Miller, C., Singh, S.S., Thomas, R., and Tayade, C. (2015). Pathophysiology and Immune Dysfunction in Endometriosis. Biomed Res. Int. 2015, 795976.

Attia, G.R., Zeitoun, K., Edwards, D., Johns, A., Carr, B.R., and Bulun, S.E. (2000). Progesterone Receptor Isoform A But Not B Is Expressed in Endometriosis1. J. Clin. Endocrinol. Metab. 85, 2897–2902.

Below, C.R., Kelly, J., Brown, A., Humphries, J.D., Hutton, C., Xu, J., Lee, B.Y., Cintas, C., Zhang, X., Hernandez-Gordillo, V., et al. (2022). A microenvironment-inspired synthetic three-dimensional model for pancreatic ductal adenocarcinoma organoids. Nat. Mater. 21, 110–119.

Blencowe, M., Arneson, D., Ding, J., Chen, Y.-W., Saleem, Z., and Yang, X. (2019). Network modeling of single-cell omics data: challenges, opportunities, and progresses. Emerg. Top. Life Sci. 3, 379–398.

Browaeys, R., Saelens, W., and Saeys, Y. (2020). NicheNet: modeling intercellular communication by linking ligands to target genes. Nat. Methods 17, 159–162.

Bulun, S.E. (2009). Endometriosis. N. Engl. J. Med. 360, 268–279.

Bulun, S.E., Yilmaz, B.D., Sison, C., Miyazaki, K., Bernardi, L., Liu, S., Kohlmeier, A., Yin, P., Milad, M., and Wei, J. (2019). Endometriosis. Endocr. Rev. 40, 1048–1079.

Burney, R.O., and Giudice, L.C. (2012). Pathogenesis and pathophysiology of endometriosis. Fertil. Steril. 98, 511–519.

Burney, R.O., Talbi, S., Hamilton, A.E., Vo, K.C., Nyegaard, M., Nezhat, C.R., Lessey, B.A., and Giudice, L.C. (2007). Gene expression analysis of endometrium reveals progesterone resistance and candidate susceptibility genes in women with endometriosis. Endocrinology 148, 3814–3826.

Chan, R.W.S., Schwab, K.E., and Gargett, C.E. (2004). Clonogenicity of Human Endometrial Epithelial and Stromal Cells1. Biol. Reprod. 70, 1738–1750.

Eskenazi, B., and Warner, M.L. (1997). Epidemiology of endometriosis. Obstet. Gynecol. Clin. North Am. 24, 235–258.

Ferrero, S. (2019). Proteomics in the Diagnosis of Endometriosis: Opportunities and Challenges. Proteomics. Clin. Appl. 13, e1800183.

Finak, G., McDavid, A., Yajima, M., Deng, J., Gersuk, V., Shalek, A.K., Slichter, C.K., Miller, H.W., McElrath, M.J., Prlic, M., et al. (2015). MAST: a flexible statistical framework for assessing transcriptional changes and characterizing heterogeneity in single-cell RNA sequencing data. Genome Biol. 16, 278.

Garcia-Alonso, L., Handfield, L.-F., Roberts, K., Nikolakopoulou, K., Fernando, R.C., Gardner, L., Woodhams, B., Arutyunyan, A., Polanski, K., Hoo, R., et al. (2021). Mapping the temporal and spatial dynamics of the human endometrium <em>in vivo</em> and <em>in vitro</em> BioRxiv 2021.01.02.425073.

Gierahn, T.M., Wadsworth, M.H., Hughes, T.K., Bryson, B.D., Butler, A., Satija, R., Fortune, S., Love, J.C., and Shalek, A.K. (2017). Seq-Well: portable, low-cost RNA sequencing of single cells at high throughput. Nat. Methods 14, 395–398.

Gnecco, J.S., Brown, A., Buttrey, K., Ives, C., Goods, B., Baugh, L., Hernandez-Gordillo, V., Loring, M., Issacson, K., and Griffith, L.G. (2021). Tissue engineered organoid co-culture model of the cycling human endometrium in a fully-defined synthetic extracellular matrix. BioRxiv 2021.09.30.462577.

Goods, B.A., Askenase, M.H., Markarian, E., Beatty, H.E., Drake, R.S., Fleming, I., DeLong, J.H., Philip, N.H., Matouk, C.C., Awad, I.A., et al. (2021). Leukocyte dynamics after intracerebral hemorrhage in a living patient reveal rapid adaptations to tissue milieu. JCI Insight 6.

Halme, J., Hammond, M.G., Hulka, J.F., Raj, S.G., and Talbert, L.M. (1984). Retrograde menstruation in healthy women and in patients with endometriosis. Obstet. Gynecol. 64, 151– 154.

Hapangama, D.K., Drury, J., Da Silva, L., Al-Lamee, H., Earp, A., Valentijn, A.J., Edirisinghe, D.P., Murray, P.A., Fazleabas, A.T., and Gargett, C.E. (2019). Abnormally located SSEA1+/SOX9+ endometrial epithelial cells with a basalis-like phenotype in the eutopic functionalis layer may play a role in the pathogenesis of endometriosis. Hum. Reprod. 34, 56– 68.

Hughes, T.K., Wadsworth, M.H., Gierahn, T.M., Do, T., Weiss, D., Andrade, P.R., Ma, F., de Andrade Silva, B.J., Shao, S., Tsoi, L.C., et al. (2020). Second-Strand Synthesis-Based Massively Parallel scRNA-Seq Reveals Cellular States and Molecular Features of Human Inflammatory Skin Pathologies. Immunity 53, 878-894.e7.

Jones, R.K., Bulmer, J.N., and Searle, R.F. (1996). Immunohistochemical characterization of stromal leukocytes in ovarian endometriosis: comparison of eutopic and ectopic endometrium with normal endometrium. Fertil. Steril. 66, 81–89.

Jørgensen, H.; Fedorcsak, P.; Isaacson, K.; Tevonian, E.; Xiao, A.; Beste, M.; Qvigstad, E.; Lauffenburger, D.; Griffith, L. (2022). Endometrial cytokines in patients with and without endometriosis evaluated for infertility. Fertil. Steril.

Käll, L., Storey, J.D., and Noble, W.S. (2008). Non-parametric estimation of posterior error probabilities associated with peptides identified by tandem mass spectrometry. Bioinformatics 24, i42–i48.

Lemke, G. (2013). Biology of the TAM receptors. Cold Spring Harb. Perspect. Biol. 5, a009076.

Lemke, G. (2019). How macrophages deal with death. Nat. Rev. Immunol. 19, 539–549.

Lemke, G., and Rothlin, C. V (2008). Immunobiology of the TAM receptors. Nat. Rev. Immunol. 8, 327–336.

Li, Q., Lu, Q., Lu, H., Tian, S., and Lu, Q. (2013). Systemic autoimmunity in TAM triple knockout mice causes inflammatory brain damage and cell death. PLoS One 8, e64812.

Lin, Y.-H., Chen, Y.-H., Chang, H.-Y., Au, H.-K., Tzeng, C.-R., and Huang, Y.-H. (2018). Chronic Niche Inflammation in Endometriosis-Associated Infertility: Current Understanding and Future Therapeutic Strategies. Int. J. Mol. Sci. 19, 2385.

Lu, Q., and Lemke, G. (2001). Homeostatic Regulation of the Immune System by Receptor Tyrosine Kinases of the Tyro 3 Family. Science (80-.). 293, 306 LP – 311.

Luecken, M.D., and Theis, F.J. (2019). Current best practices in single-cell RNA-seq analysis: a tutorial. Mol. Syst. Biol. 15, e8746.

McGinnis, C.S., Murrow, L.M., and Gartner, Z.J. (2019). DoubletFinder: doublet detection in single-cell RNA sequencing data using artificial nearest neighbors. Cell Syst. 8, 329–337.

McKinnon, B., Mueller, M., and Montgomery, G. (2018). Progesterone Resistance in Endometriosis: an Acquired Propertyã Trends Endocrinol. Metab. 29, 535–548.

McQuin, C., Goodman, A., Chernyshev, V., Kamentsky, L., Cimini, B.A., Karhohs, K.W., Doan, M., Ding, L., Rafelski, S.M., Thirstrup, D., et al. (2018). CellProfiler 3.0: Next-generation image processing for biology. PLoS Biol. 16, e2005970.

van der Meer, J.H.M., van der Poll, T., and van ‘t Veer, C. (2014). TAM receptors, Gas6, and protein S: roles in inflammation and hemostasis. Blood 123, 2460–2469.

Meuleman, C., Vandenabeele, B., Fieuws, S., Spiessens, C., Timmerman, D., and D’Hooghe, T. (2009). High prevalence of endometriosis in infertile women with normal ovulation and normospermic partners. Fertil. Steril. 92, 68–74.

Moore, L., Leongamornlert, D., Coorens, T.H.H., Sanders, M.A., Ellis, P., Dentro, S.C., Dawson, K.J., Butler, T., Rahbari, R., Mitchell, T.J., et al. (2020). The mutational landscape of normal human endometrial epithelium. Nature 580, 640–646.

Nagata, K., Ohashi, K., Nakano, T., Arita, H., Zong, C., Hanafusa, H., and Mizuno, K. (1996). Identification of the Product of Growth Arrest-specific Gene 6 as a Common Ligand for Axl, Sky, and Mer Receptor Tyrosine Kinases*. J. Biol. Chem. 271, 30022–30027.

Nakamura, Y.S., Hakeda, Y., Takakura, N., Kameda, T., Hamaguchi, I., Miyamoto, T., Kakudo, S., Nakano, T., Kumegawa, M., and Suda, T. (1998). Tyro 3 Receptor Tyrosine Kinase and its Ligand, Gas6, Stimulate the Function of Osteoclasts. Stem Cells 16, 229–238.

Nesvizhskii, A.I., Keller, A., Kolker, E., and Aebersold, R. (2003). A statistical model for identifying proteins by tandem mass spectrometry. Anal. Chem. 75, 4646–4658.

Nnoaham, K.E., Hummelshoj, L., Webster, P., d’Hooghe, T., de Cicco Nardone, F., de Cicco Nardone, C., Jenkinson, C., Kennedy, S.H., and Zondervan, K.T. (2011). Impact of endometriosis on quality of life and work productivity: a multicenter study across ten countries. Fertil. Steril. 96, 366-373.e8.

Oberg, A.L., Mahoney, D.W., Eckel-Passow, J.E., Malone, C.J., Wolfinger, R.D., Hill, E.G., Cooper, L.T., Onuma, O.K., Spiro, C., Therneau, T.M., et al. (2008). Statistical analysis of relative labeled mass spectrometry data from complex samples using ANOVA. J. Proteome Res. 7, 225–233.

Ordovas-Montanes, J., Beyaz, S., Rakoff-Nahoum, S., and Shalek, A.K. (2020). Distribution and storage of inflammatory memory in barrier tissues. Nat. Rev. Immunol. 20, 308–320.

Osteen, K.G., Hill, G.A., Hargrove, J.T., and Gorstein, F. (1989). Development of a method to isolate and culture highly purified populations of stromal and epithelial cells from human endometrial biopsy specimens. Fertil. Steril. 52, 965–972.

Prašnikar, E., Knez, J., Kovačič, B., and Kunej, T. (2020). Molecular signature of eutopic endometrium in endometriosis based on the multi-omics integrative synthesis. J. Assist. Reprod. Genet. 1–19.

Qu, H., Li, L., Wang, T.-L., Seckin, T., Segars, J., and Shih, I.-M. (2020). Epithelial Cells in Endometriosis and Adenomyosis Upregulate STING Expression. Reprod. Sci. 27, 1276–1284.

Rai, P., Kota, V., Sundaram, C.S., Deendayal, M., and Shivaji, S. (2010). Proteome of human endometrium: Identification of differentially expressed proteins in proliferative and secretory phase endometrium. PROTEOMICS – Clin. Appl. 4, 48–59.

Raudvere, U., Kolberg, L., Kuzmin, I., Arak, T., Adler, P., Peterson, H., and Vilo, J. (2019). g:Profiler: a web server for functional enrichment analysis and conversions of gene lists (2019 update). Nucleic Acids Res. 47, W191–W198.

Sampson, J.A. (1918). The escape of foreign material from the uterine cavity into the uterine veins (William Wood & Company).

Shadforth, I.P., Dunkley, T.P.J., Lilley, K.S., and Bessant, C. (2005). i-Tracker: For quantitative proteomics using iTRAQ™. BMC Genomics 6, 145.

Shigesi, N., Kvaskoff, M., Kirtley, S., Feng, Q., Fang, H., Knight, J.C., Missmer, S.A., Rahmioglu, N., Zondervan, K.T., and Becker, C.M. (2019). The association between endometriosis and autoimmune diseases: a systematic review and meta-analysis. Hum. Reprod. Update 25, 486–503.

Simoens, S., Dunselman, G., Dirksen, C., Hummelshoj, L., Bokor, A., Brandes, I., Brodszky, V., Canis, M., Colombo, G.L., DeLeire, T., et al. (2012). The burden of endometriosis: costs and quality of life of women with endometriosis and treated in referral centres. Hum. Reprod. 27, 1292–1299.

Simpson, J.L., Elias, S., Malinak, L.R., and Buttram Jr, V.C. (1980). Heritable aspects of endometriosis: I. Genetic studies. Am. J. Obstet. Gynecol. 137, 327–331.

Symons, L.K., Miller, J.E., Kay, V.R., Marks, R.M., Liblik, K., Koti, M., and Tayade, C. (2018). The immunopathophysiology of endometriosis. Trends Mol. Med. 24, 748–762.

Tan, Y., Flynn, W.F., Sivajothi, S., Luo, D., Bozal, S.B., Luciano, A.A., Robson, P., Luciano, D.E., and Courtois, E.T. (2021). Single cell analysis of endometriosis reveals a coordinated transcriptional program driving immunotolerance and angiogenesis across eutopic and ectopic tissues. BioRxiv 2021.07.28.453839.

Tapmeier, T.T., Rahmioglu, N., Lin, J., De Leo, B., Obendorf, M., Raveendran, M., Fischer, O.M., Bafligil, C., Guo, M., Harris, R.A., et al. (2021). Neuropeptide S receptor 1 is a nonhormonal treatment target in endometriosis. Sci. Transl. Med. 13.

Vago, J.P., Amaral, F.A., and van de Loo, F.A.J. (2021). Resolving inflammation by TAM receptor activation. Pharmacol. Ther. 227, 107893.

Vallvé-Juanico, J., Houshdaran, S., and Giudice, L.C. (2019). The endometrial immune environment of women with endometriosis. Hum. Reprod. Update 25, 565–592.

Vogel, C., and Marcotte, E.M. (2012). Insights into the regulation of protein abundance from proteomic and transcriptomic analyses. Nat. Rev. Genet. 13, 227–232.

Vullings, J., Vago, J.P., Waterborg, C.E.J., Thurlings, R.M., Koenders, M.I., van Lent, P.L.E.M., van der Kraan, P.M., Amaral, F.A., and van de Loo, F.A.J. (2020). Selective Increment of Synovial Soluble TYRO3 Correlates with Disease Severity and Joint Inflammation in Patients with Rheumatoid Arthritis. J. Immunol. Res. 2020, 9690832.

Wang, W., Vilella, F., Alama, P., Moreno, I., Mignardi, M., Isakova, A., Pan, W., Simon, C., and Quake, S.R. (2020). Single-cell transcriptomic atlas of the human endometrium during the menstrual cycle. Nat. Med. 26, 1644–1653.

Warren, L.A., Shih, A., Renteira, S.M., Seckin, T., Blau, B., Simpfendorfer, K., Lee, A., Metz, C.N., and Gregersen, P.K. (2018). Analysis of menstrual effluent: diagnostic potential for endometriosis. Mol. Med. 24, 1.

Weng, L., Hou, S., Lei, S., Peng, H., Li, M., and Zhao, D. (2020). Estrogen-regulated CD200 inhibits macrophage phagocytosis in endometriosis. J. Reprod. Immunol. 138, 103090.

Xie, Q., He, H., Wu, Y.-H., Zou, L.-J., She, X.-L., Xia, X.-M., and Wu, X.-Q. (2019). Eutopic endometrium from patients with endometriosis modulates the expression of CD36 and SIRP-α in peritoneal macrophages. J. Obstet. Gynaecol. Res. 45, 1045–1057.

Zondervan, K.T., Becker, C.M., and Missmer, S.A. (2020). Endometriosis. N. Engl. J. Med. 382, 1244–1256.

