## Supplementary figures and images for "Integrating endometrial proteomic and single cell transcriptomic pipelines reveals distinct menstrual cycle and endometriosis-associated molecular profiles"

### Supplemental Figure 1

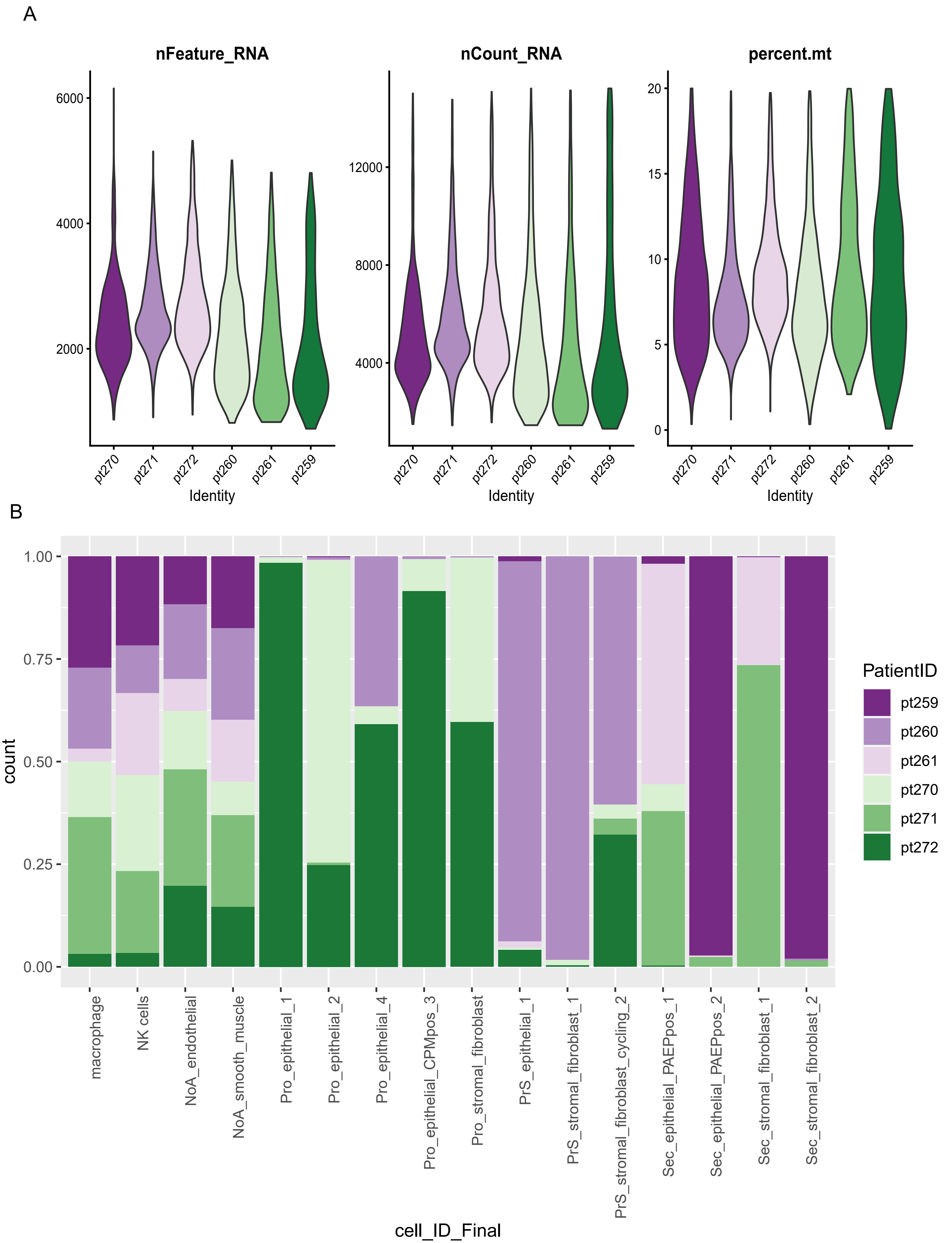

### Supplemental Figure 2

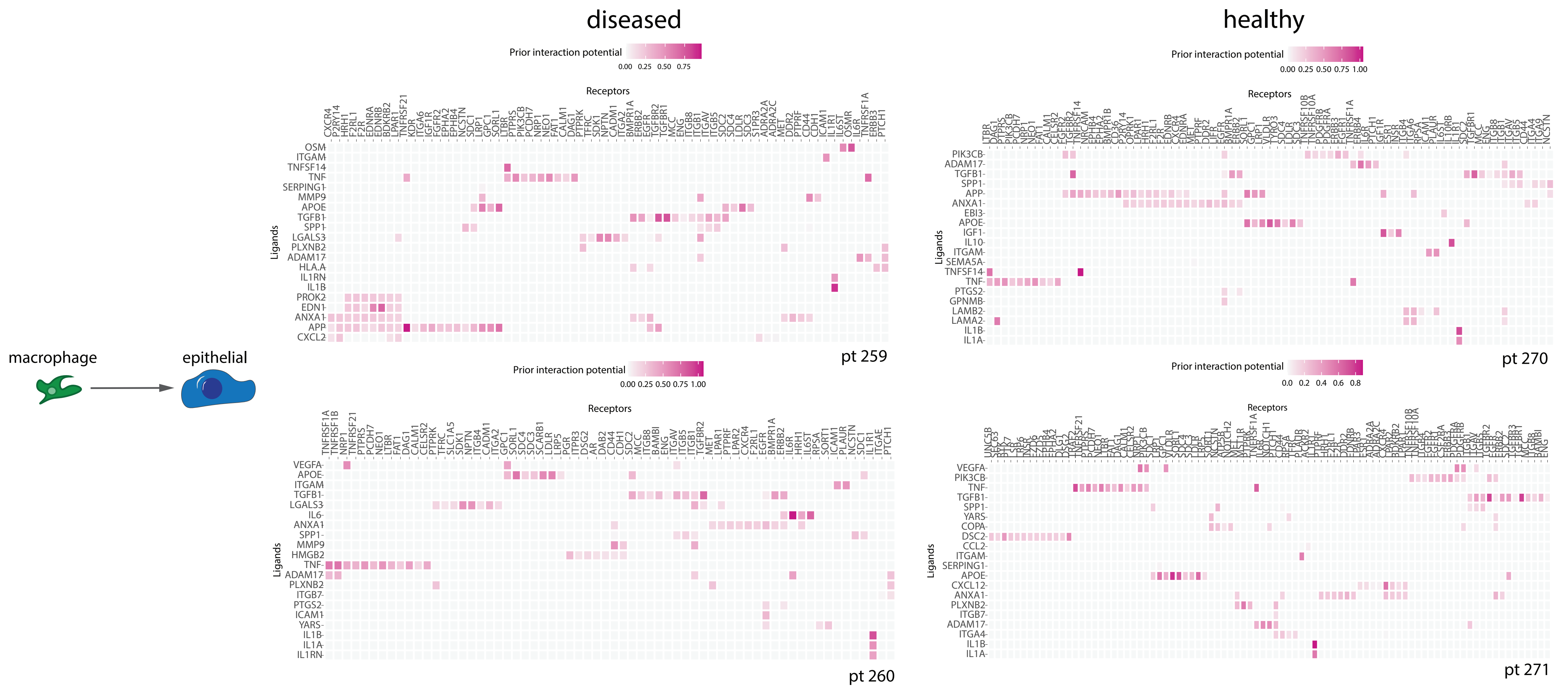
