## Supplemental Figure 3 for "Integrating endometrial proteomic and single cell transcriptomic pipelines reveals distinct menstrual cycle and endometriosis-associated molecular profiles"

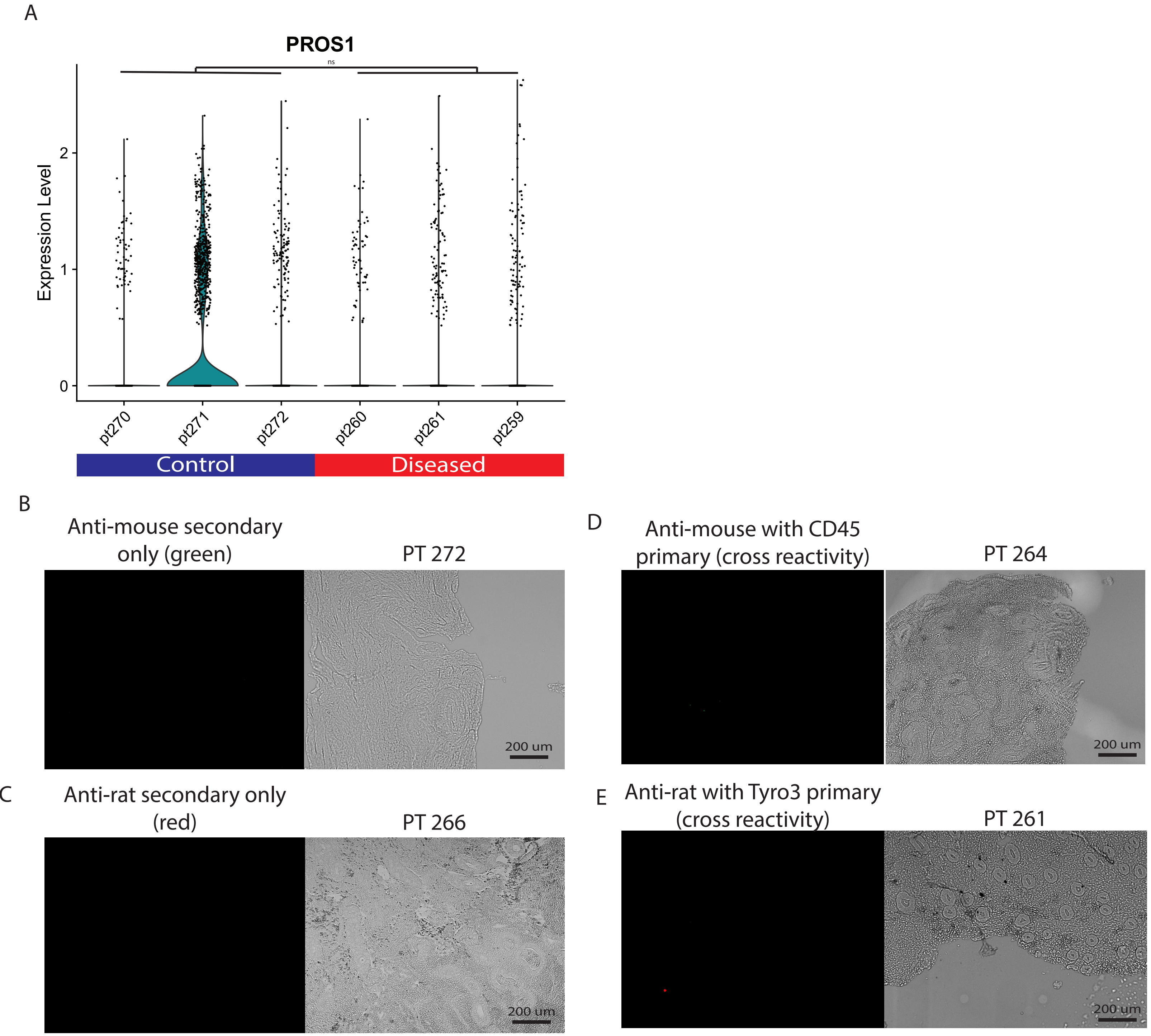


Supplemental Figure 2. Expression of Protien S and seondary IHC controls. A. Expression of Protein S (PROS1) mirrors Tryo3 expression across all patient cells measured with single cells RNA sequencing. B. Control staining with no primary and the anti-mouse secondary only. C. Control staining with no primary and anti-rat secondary only. D. Control staining for cross reactivity with CD45 (rat host) as the primary and anti-mouse secondary. E. Control staining for cross reactivity with Tryo3 (mouse host) as the primary and anti-rat secondary.
